# Developing a psychological support intervention to help injured athletes get Back in the Game

**DOI:** 10.1101/2021.03.01.21252681

**Authors:** Clare L. Ardern, Nicholas Hooper, Paul O’Halloran, Kate E. Webster, Joanna Kvist

## Abstract

**Background:** After serious knee injury, up to half of athletes do not return to competitive sport, despite recovering sufficient physical function. Athletes often desire psychological support to return to sport, but rehabilitation clinicians feel ill-equipped to deliver adequate support.

**Objective:** To design and develop an Internet-delivered psychological support programme for athletes recovering from knee ligament surgery.

**Method:** Our work developing and designing the Back in the Game intervention was guided by a blend of *theory & evidence-based* and *target population-based* strategies to developing complex interventions. We systematically searched for qualitative evidence related to athletes’ experiences, perspectives and needs for recovery and return to sport after anterior cruciate ligament (ACL) injury. Two reviewers coded and synthesised the results using thematic meta-synthesis. We systematically searched for randomised controlled trials (RCTs) reporting on psychological support interventions for improving ACL rehabilitation outcomes in athletes. One reviewer extracted the data (including effect estimates); a second reviewer checked the data for accuracy. The results were synthesised descriptively. We conducted feasibility testing in two phases: (1) technical assessment, and (2) feasibility and useability testing. For phase 1, we recruited clinicians and people with lived experience of ACL injury. For phase 2, we recruited patients aged between 15 and 30 years, who were within 8 weeks of ACL reconstruction surgery. Participants completed a 10-week version of the intervention, and semi-structured interviews evaluating acceptability, demand, practicality and integration. The project was approved by the Swedish Ethical Review Authority (2018/45-31).

**Results:** Three analytic themes emerged from the meta-synthesis (*n* = 16 studies, 164 participants): (1) tools/strategies to support rehabilitation progress, (2) barriers and facilitators for physical readiness to return to sport, and (3) barriers and facilitators to psychological readiness to return to sport. Coping strategies, relaxation and goal setting may have a positive effect on rehabilitation outcomes after ACL reconstruction (*n* = 7 RCTs, 430 participants). There were no trials of psychological support interventions for improving return to sport. Eleven people completed phase 1 of feasibility testing (technical assessment) and identified 4 types of software errors that we fixed. Six participants completed feasibility and useability testing. Their feedback suggested the intervention was easy to access and addressed the needs of athletes who want to return to sport after ACL reconstruction. We refined the intervention to include more multimedia content, and support to access and use the intervention features.

**Conclusion:** The Back in the Game intervention is a 24-week Internet-delivered self-guided programme comprising 7 modules that complements usual rehabilitation, changes focus as rehabilitation progresses, is easy to access and use, and includes different psychological support strategies.

## Background

This paper describes the processes for developing an Internet-delivered programme providing psychological support for return to sport to athletes recovering from knee ligament surgery (anterior cruciate ligament (ACL) reconstruction). Development is the entire process to arrive at an intervention to test in a randomised controlled trial. Design is the specific decisions we made about intervention content, format and delivery mode.[53]

We adopted a blended approach[52, 53] to developing and designing the Back in the Game intervention, which leaned heavily on *theory & evidence-based* (UK Medical Research Council Framework for developing and evaluating complex interventions[27]) and *target population-based* (person-based [73]) approaches. O’Cathain’s et al. taxonomy and synthesis[53] guided developing the 8 ‘building blocks’ that underpin our work. This paper is structured around the 8 building blocks (Figure 1).

**Figure 1.**
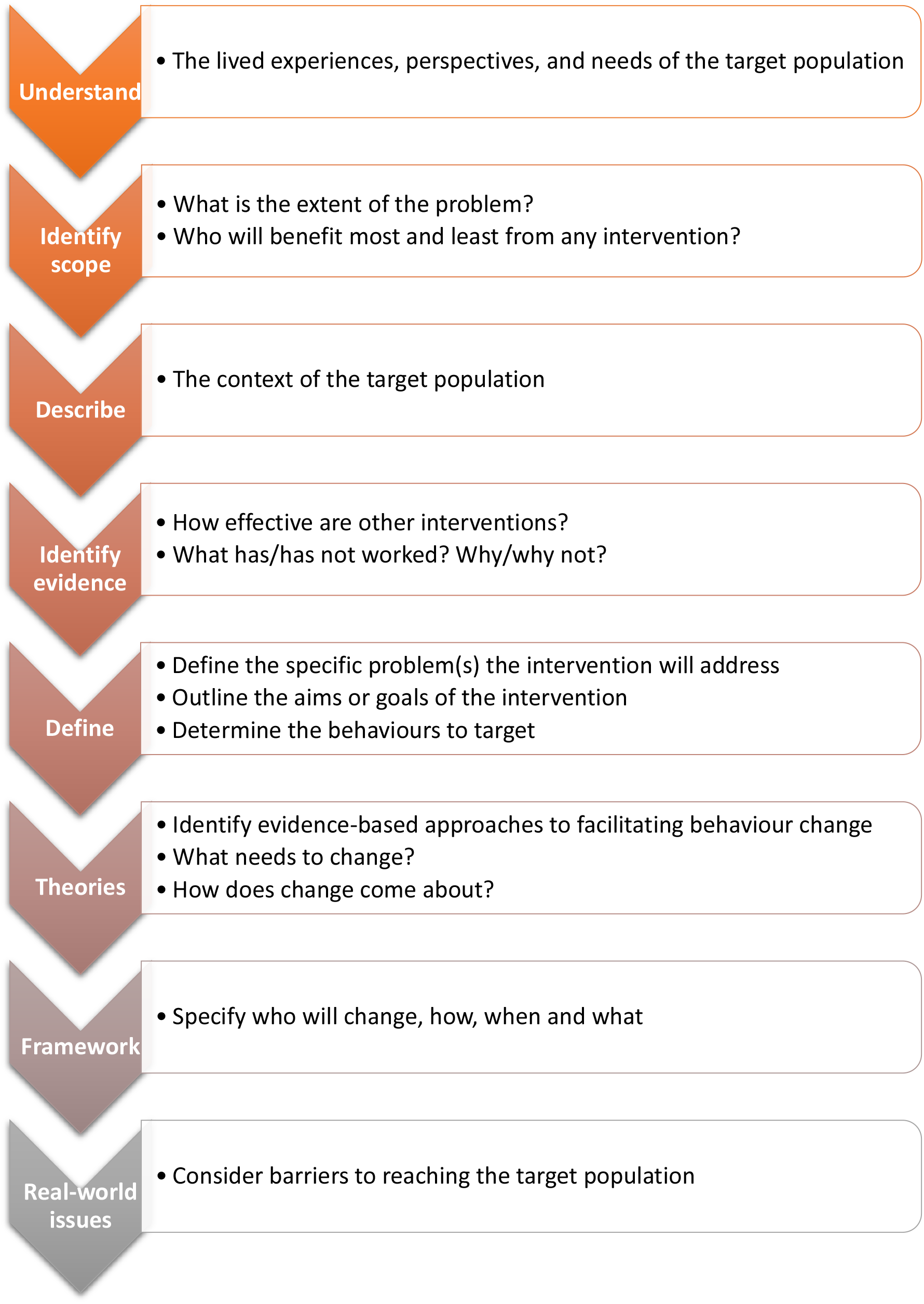
Eight building blocks underpin our work to develop the Back in the Game intervention.

### Building block 1: Understand the problems or issues to be addressed, based on the lived experience, perspectives and needs of active people with ACL reconstruction

We systematically searched electronic databases to identify qualitative research exploring the perceptions and experience of active people with ACL reconstruction. We used the PerSPecTIF framework[17] to frame the research question for a qualitative evidence synthesis: *From the perspective of an active person with ACL reconstruction, in the setting of completing or having completed rehabilitation, how does the phenomenon of biopsychosocial factors during recovery impact on a person’s experiences and perceptions related to recovery and return to sport?* The meta-synthesis methods and results from 16 qualitative studies are outlined in Appendix 1. Sixteen descriptive themes emerged, which we mapped to 3 analytic themes to inform developing the Back in the Game intervention: (1) barriers and facilitators for *psychological readiness to return to sport*, (2) barriers and facilitators for *physical readiness to return to sport*, and (3) *tools or strategies* to support rehabilitation progress.

### Evidence synthesis

Athletes at all levels (amateur to professional) shared common perceptions and lived experience of rehabilitation after ACL reconstruction. Athletes wanted to play sport and saw playing sport as central to their self-identity. They felt happy when they were playing sport. Although, the experience of ACL injury had often irrevocably changed how they thought of themselves and their capacity to contribute to society.

Anxiety and low confidence were dominant emotions. Athletes felt scared, uncertain, frustrated, and hopeless at different times during recovery and when returning to sport. Sometimes athletes avoided tasks or activities (e.g. sport-specific movements) because they lacked confidence in their knee. Athletes drew support, feedback, encouragement and reassurance from people they trusted. Previous experience of sports injury also helped athletes know what was required to recover and return to sport.

Athletes judged that quality rehabilitation programmes included strategies to help build their physical and mental capacity to participate safely in sport again. Highlight box 1 summarises the key aspects of athletes’ perspectives, lived experience of rehabilitation and needs after ACL reconstruction that informed developing and designing the intervention (see appendix 1, table A1b for detailed summary).

### Highlight box 1

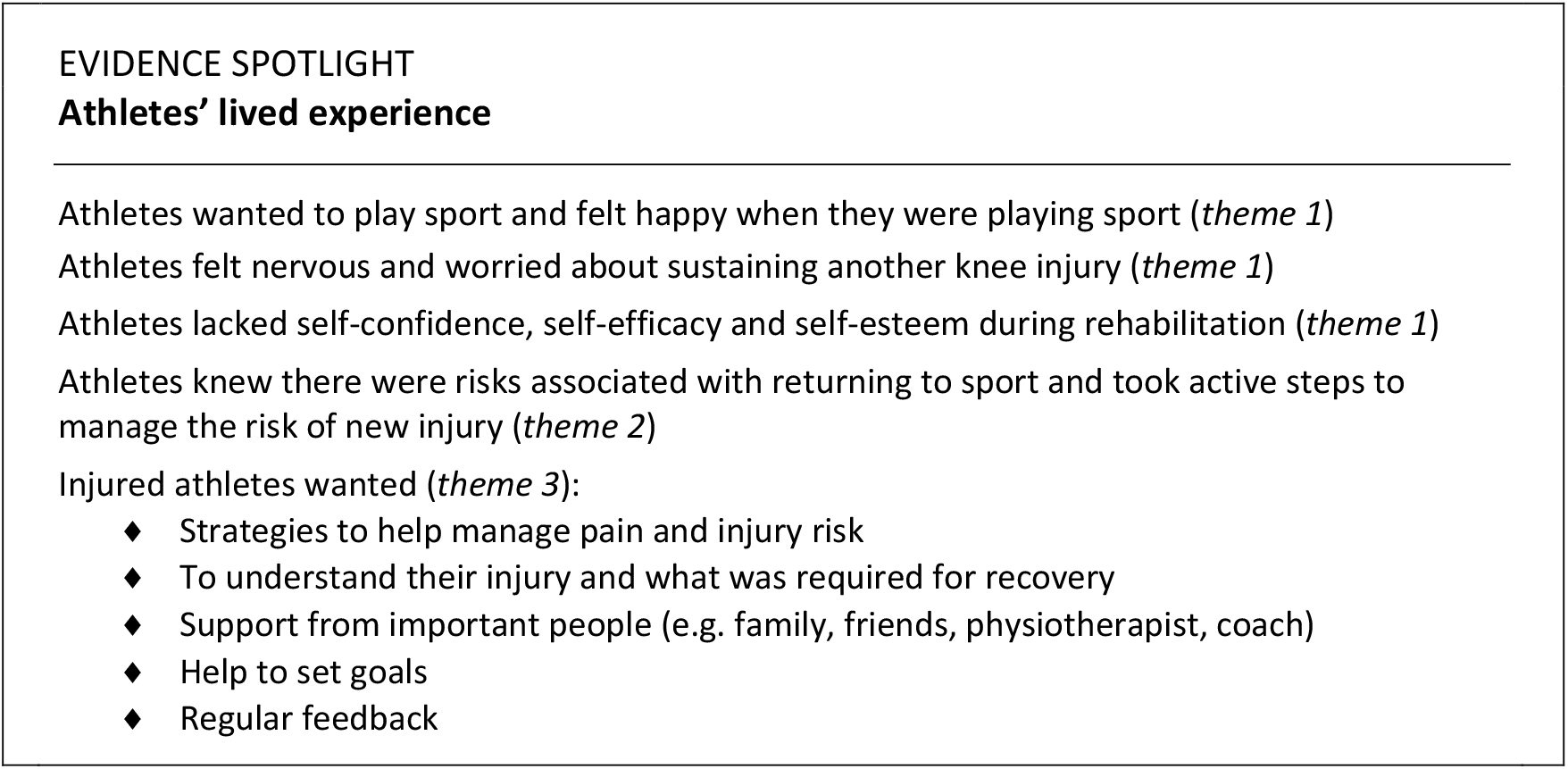

### Building block 2: Identify the scope of the problem

After serious knee injury, up to half of athletes do not return to competitive sport.[11] Athletes typically recover sufficient knee function to withstand the physical demands of playing their sport, but the athlete’s mental state is often the main hurdle to returning[13]— physical and psychological readiness to return to sport often do not coincide. Fear of reinjury is the most frequent reason reported by athletes who do not return to their preinjury sport, or give up sport after ACL injury or reconstruction.[10, 13] Athletes of all ages at all participation levels can have problems returning to sport after ACL reconstruction.[11, 43]

### What contributes to the problem?

Young athletes toiling in rehabilitation get bored and lose motivation during the long rehabilitation period.[29, 55] They feel frustrated when rehabilitation does not focus on sports performance,[14, 15] and concerned about their body’s ability to cope with the demands of their sport.[57] Anxiety about the consequences of sustaining the knee injury again often besets athletes as they work towards returning to sport.[56]

Psychological factors have large effects on return to sport outcomes after ACL reconstruction, and larger effects on outcomes than physical factors.[6, 72] Greater psychological readiness (a construct incorporating confidence, emotions and risk appraisal) is associated with greater likelihood of returning to the preinjury sport.[11]

### Who benefits from a psychological support intervention?

Athletes with ACL injury expect and desire to return to sport.[40, 42, 71] Up to 9 in every 10 recreational athletes expect to return to their preinjury sport after ACL reconstruction.[33, 71] Yet, fewer athletes than expected achieve their return to sport goals.[11] Psychological factors (including confidence, anxiety and risk appraisal), many of which are potentially modifiable, exert a strong influence on athletes achieving their return to sport goals.[6, 11] Therefore, an effective intervention providing psychological support for return to sport is likely to be relevant for a majority of athletes. Although, the intervention may not specifically benefit athletes who have high confidence and motivation to return to sport (and maintain their confidence and motivation through rehabilitation), or athletes who dislike psychological support.

### Building block 3: Describe the context of the target population

A biopsychosocial approach to health and rehabilitation is the dominant paradigm within which health care is delivered in the 21st Century.[31] Clinicians recognise that different patients require different emphasis on biological, psychological, and social elements, and different emphasis at different times during a course of treatment. The transition through rehabilitation and resuming sport after injury can be difficult. To add to the challenge, athletes are often discharged from rehabilitation months before attempting to return to sport. Critically, most athletes lack the support of a rehabilitation clinician during the transition back to sport.

### Managing anterior cruciate ligament injury in the 2020s: usual rehabilitation for ACL injury

High-quality rehabilitation aims to help athletes gradually regain knee function and prepare to return to sport. Recovering physical function is vital for achieving return to sport goals— athletes require sufficient physical capacity to cope with the demands of playing sport, to execute their skills as desired, and stay injury-free.[39]

Rehabilitation clinicians foster a positive rehabilitation environment where the athlete is appropriately physically and mentally challenged using regular monitoring, assessment and feedback to maintain or build physical function, motivation and confidence.[36-38, 67] The rehabilitation clinician is the conductor of a multidisciplinary ‘orchestra’–connecting clinicians, coaches, the athlete, parents, and others who may all contribute to shared decision-making at different points during rehabilitation. The ultimate prize: safe and timely return to sport.

The treating clinician and the athlete collaborate to decide on specific therapies and exercises, and the number of face-to-face, home-based and gymnasium-based training sessions required to achieve the rehabilitation aims. The volume and content of usual post-operative rehabilitation is highly variable, although, clinical practice guidelines recommend at least 3 sessions per week. Rehabilitation programmes typically run for at least 6 months, and usually cease by 12 months.

### Four rehabilitation phases for return to pivoting sport[8]

1. **Acute phase** aimed at reducing pain and swelling, improving knee movement, and recovering performance of activities of daily living (e.g. walking without aids).
2. **Intermediate phase** aimed at progressing knee function and muscle strength in more demanding and sport-specific tasks–emphasising dynamic knee stability.
3. **Late phase** aimed at gradually transitioning from gym-based exercises to the training and match load demands of the athlete’s sport. Return to unrestricted participation in the athlete’s goal sport is the end of late phase rehabilitation. There is staged progression from modified training (participation in non-contact activities only), to full training (no restrictions on contact), to restricted match play (e.g. limited time in the game), to unrestricted match play.
4. **Injury prevention phase** aimed at embedding exercises focused on movement control into the athlete’s usual training and match preparation. A programme emphasising muscle strength and dynamic knee stability should be performed at least twice per week as part of normal training. The programme must be combined with appropriate workload management to minimise the risk for new injuries to other structures and reinjury to the knee.

Return to sport occurs along a continuum,[7] beginning at injury diagnosis and concluding when the athlete is performing as desired in their chosen sport. Therefore, return to sport support should also be delivered along the same continuum.

### Building block 4: Identify evidence of effectiveness of other psychological support interventions

We systematically searched for research addressing the question: “*What is the efficacy of psychological support interventions for improving ACL injury rehabilitation outcomes in athletes*?”. We identified and qualitatively summarised the major characteristics of psychological support for athletes and the consequences of providing psychological support during rehabilitation after ACL reconstruction. The aim was to articulate a credible causal explanation for a psychological support intervention improving return to sport after ACL reconstruction. The review methods, and specific results and quality assessment from the 7 included randomised controlled trials are outlined in Appendix 2.

### Evidence summary

Psychological skills training that targets coping strategies, relaxation and goal setting might work for improving physical impairments and psychological outcomes after ACL reconstruction. It is uncertain whether imagery/visualisation is helpful for the injured athlete. Highlight box 2 outlines treatment approaches with potential to improve health outcomes to include in a new psychological support intervention.

### Highlight box 2

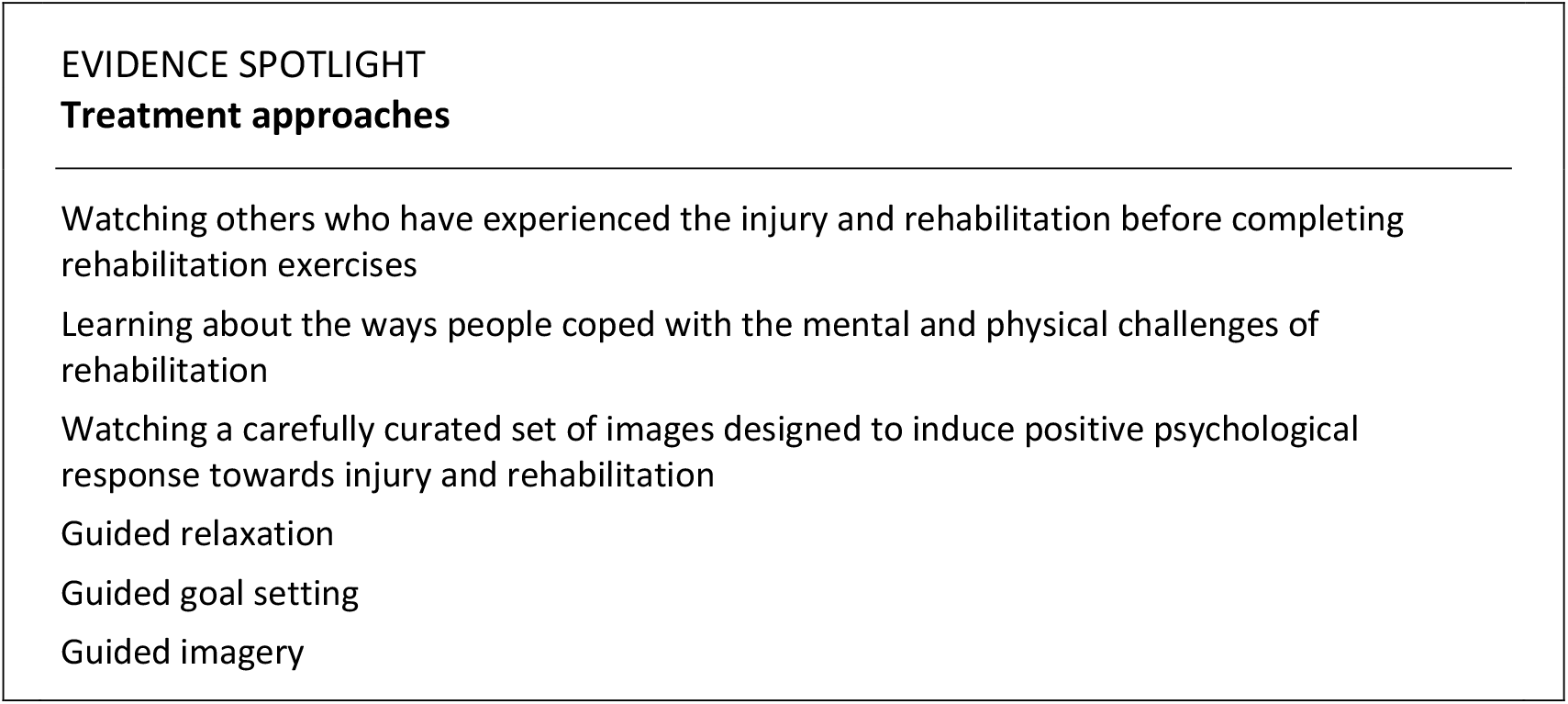

### Building block 5: Define the problem and identify behaviours to target

The mind matters for returning to sport.[10, 12] The clinician may frequently encounter the athlete who has the physical capabilities to participate in sport, but is anxious about participating.[13] Athletes desire psychological support during rehabilitation, [15, 61] yet many clinicians feel underequipped to provide effective psychological support.[5, 30] Conspicuous by its absence in clinical practice guidelines,[1, 70] is a systematic approach to addressing psychological responses throughout a rehabilitation programme. While experienced clinicians may provide tacit psychological support, there is no widely available, evidence-based programme for injured athletes.

### Aim and goal of the Back in the Game intervention

To deliver on-demand psychological support in parallel with usual post-operative rehabilitation. The goal is to improve athletes’ confidence to return to sport, and subsequently improve the return to preinjury sport rate after ACL reconstruction.

### Highlight box 3

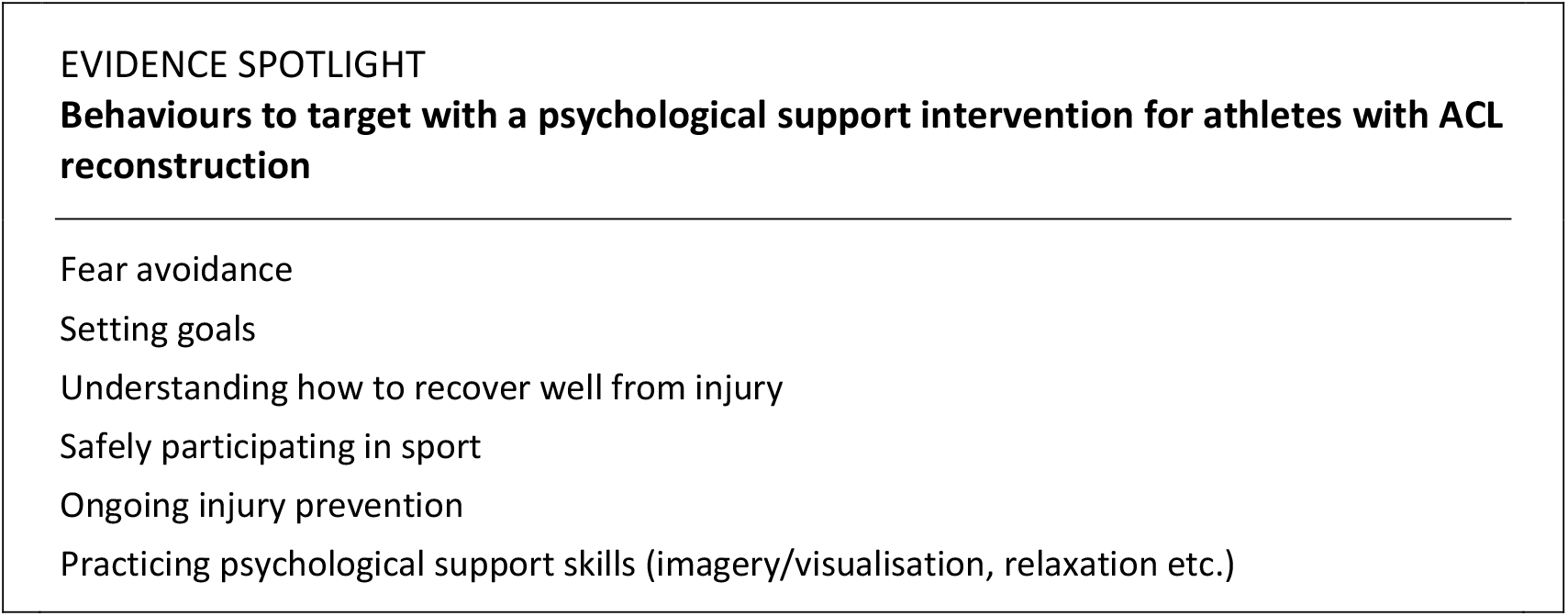

### Behaviours to target

A psychological support intervention to help athletes return to sport after injury should deliver practical tools or strategies that athletes can use to complement physical rehabilitation (highlight box 3). The intervention should (i) help injured athletes understand their injury and what is required for recovery, (ii) support athletes to set realistic goals, and provide regular feedback on progress towards athletes’ goals, (iii) teach athletes strategies to manage fear and anxiety, boost low confidence and self-efficacy, and maintain motivation and athlete identity, and (iv) support athletes to establish life-long habits and stay healthy while playing sport.

### Building block 6: Theories—ways to facilitate return to sport behaviour change

The Back in the Game intervention is aimed at promoting behaviour change—from not participating in sport due to injury, to playing the preinjury sport. During rehabilitation, athletes with ACL injury are not participating in their desired sport, and the aim of the intervention is to support athletes to reach their desired sports participation (behaviour) goal. The intervention is grounded in cognitive theory and self-determination theory, and motivational interviewing.

According to cognitive theory, how a person believes, perceives, plans and interprets their world influences, and is influenced by, their behaviour and emotions.[19, 44] Cognitive theory underpins cognitive-behavioural therapy. The aim of cognitive behavioural therapy is to transform negative thoughts/thinking to positive thoughts/thinking by changing a person’s thoughts, emotions and behaviours. Cognitive-behavioural therapy targets a person’s interpretations and beliefs. Uncovering and changing negative thinking patterns boosts the person’s self-motivation to engage in a health-promoting behaviour.[19]

We recognised that motivation resonated in athletes’ lived experience of recovering from ACL reconstruction, and incorporated self-determination theory into our framework for Back in the Game. Motivation to change one’s behaviour is strongest when one feels the behaviour is self-determined.[58] For return to sport, an athlete’s self-motivation to engage in sport (i.e. fulfil the return to sport goals) is driven by three key elements: a sense of personal control over what happens (autonomy), a belief that one has the skills and knowledge to succeed (competence), and support to achieve one’s goals (relatedness).[58]

Combining cognitive theory and self-determination theory, we propose that when one has positive thoughts about the behaviour, motivation to engage in the target behaviour is enhanced, ultimately boosting the likelihood of engaging in and sustaining the behaviour. Using cognitive behavioural therapy to change negative thinking about one’s ability and capacity to participate in sport to positive thinking about one’s ability boosts self-motivation to engage in the behaviour, and a self-motivated person changes their behaviour (Figure 2).

**Figure 2.**
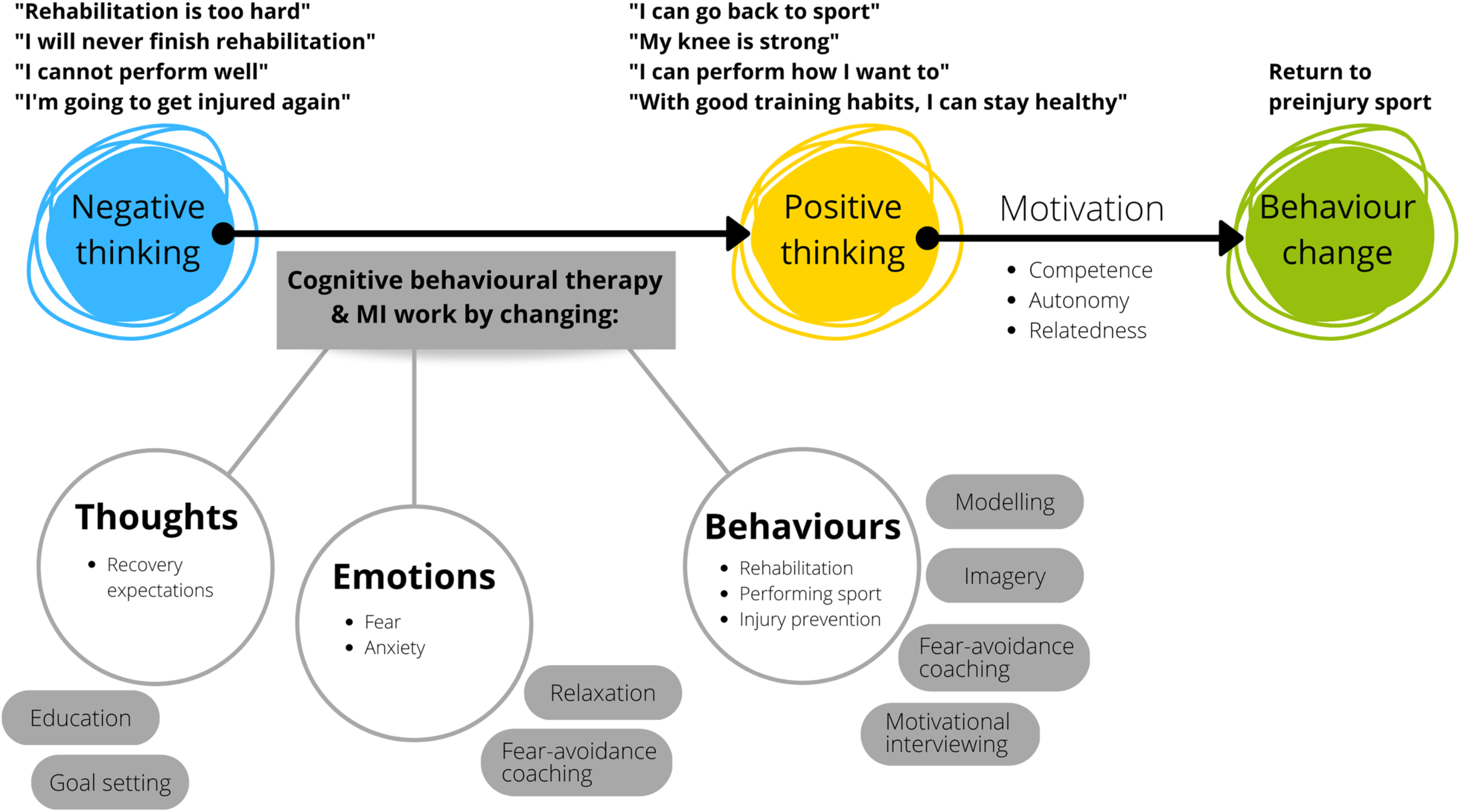
Back in the Game is grounded in cognitive theory and self-determination theory, and aims to facilitate return to sport behaviour change. Topics listed in grey ovals adjacent to thoughts, emotions and behaviours are the key contents of the intervention modules. (*Note*. MI; motivational interviewing)

Motivational interviewing emphasises core components of cognitive theory (e.g. developing self-efficacy and respecting autonomy, which are inherent to self-determination theory). Motivational interviewing strategies also facilitate motivation by drawing on the individual’s own reasons and ideas for change.[50]

### Building block 7: Framework for getting Back in the Game—who, how, when and what?

A person-based approach[73] informed the decisions we made about intervention content, format and delivery mode (designing the intervention). The approach aims for designers to understand what to do to ensure an attractive intervention that (i) addresses end users’ needs, a*nd* (ii) is feasible to implement.[73] When designing an intervention, the designer seeks to understand how different people in different situations may view and engage with an intervention. The designer may pose questions like “which elements of the intervention are helpful for this particular person?” and “which elements could be taken out of the intervention?”.

### Guiding principles for designing the Back in the Game intervention

We established 5 guiding principles[73] for intervention design (Highlight box 4). The guiding principles addressed key objectives of intervention design, and important features that must be addressed to achieve the intervention objectives.[73]

### Highlight box 4

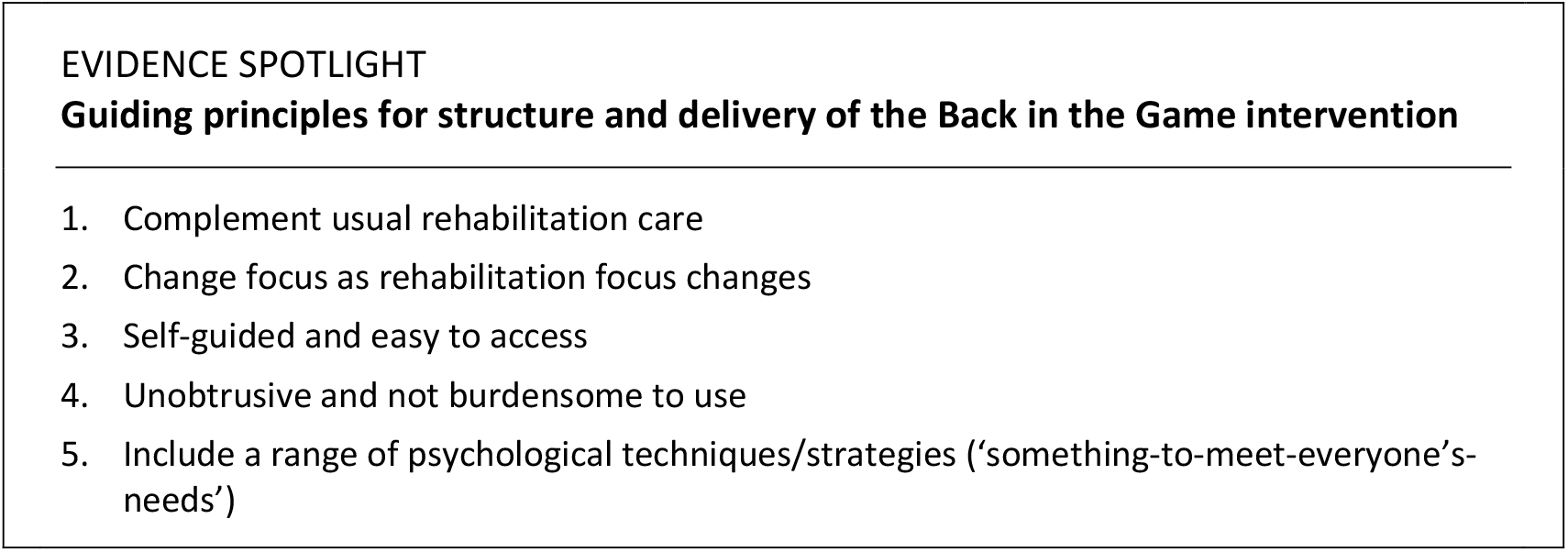

### Who will change and how will behaviour change occur?

Back in the Game delivers cognitive behavioural therapy plus motivational interviewing to help athletes identify negative thoughts about the behaviour of playing sport, and reframe the negative thoughts to positive thoughts and behaviour change. Quality cognitive-behavioural therapy is more than psychotherapy alone—it teaches psychological skills including problem solving, mindfulness and relaxation, exposure, role play, and imagery.[16] Motivational interviewing draws on the person’s own motives and ideas for change.[50] The benefit of a multifaceted approach is that it helps the athlete learn and practice a range of psychological skills. Targeting multiple skills might widen the scope of possible therapeutic benefits, and increase the likelihood of finding a skill that appeals to the athlete.

The rationale for the content and delivery is that supporting athletes to reach psychological readiness to return to sport while they complete usual post-operative rehabilitation, helps them make a more successful transition back to the preinjury sport.

### When will behaviour change occur?

Back in the Game is a 24-week intervention that commences in the first week following ACL reconstruction. Seven modules (Figure 3) are delivered in parallel with usual rehabilitation care. The intervention mirrors the progression of rehabilitation, functions as a stand-alone eHealth intervention, and does not require monitoring or input from the clinician responsible for delivering rehabilitation. Tasks are delivered in a progressive fashion, tailored to the stage of face-to-face rehabilitation. Athletes choose, from a menu of different tasks, the task that best suits their needs during each intervention session.

**Figure 3.**
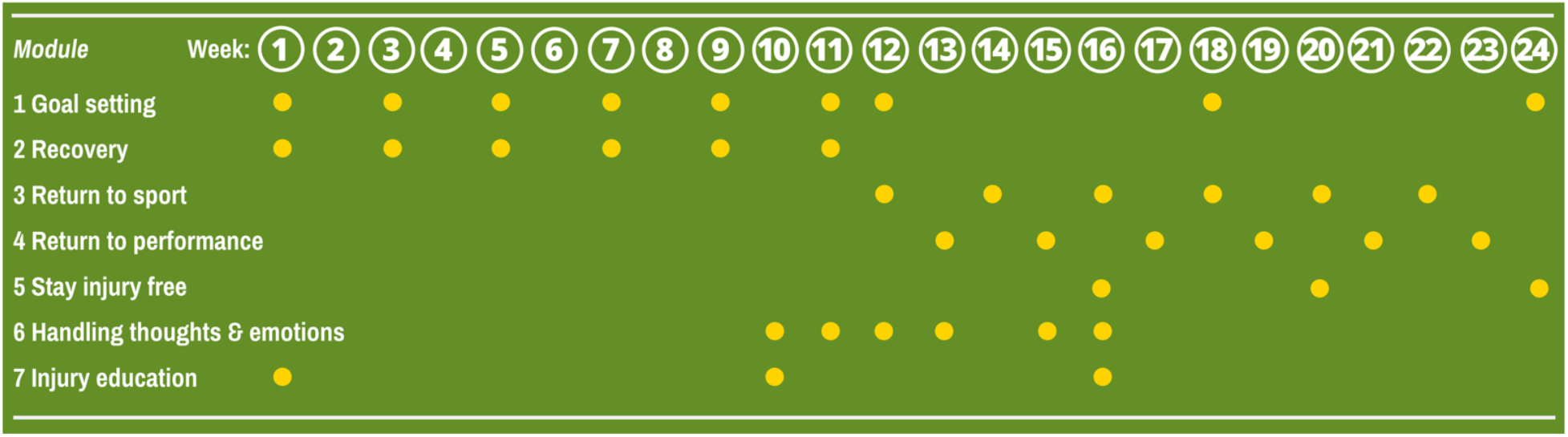
Summary of the 7 self-directed modules (covering psychological skills, psychoeducation and principles of motivational interviewing) of the Back in the Game intervention. Each dot represents how often the user is prompted to complete a task in each module.

We recommend athletes use the intervention for a minimum of 30 minutes every week. It is rare for athletes to return to pivoting sport before 6 months post-operative, and athletes are actively encouraged to delay returning to unrestricted participation in pivoting sport for at least 9 months.[39] The intervention is designed to help athletes *prepare* to return to sport; we do not expect athletes to return to their preinjury sport during the 24-week intervention period. However, we expect some athletes will be participating in modified sport (e.g. no contact, no direction changes) by the end of the intervention period.

### What will foster behaviour change?

Back in the Game employs a mix of psychological skills, psychoeducation and motivational interviewing (Figure 2). Our blended approach to developing the intervention ensured the content choices were informed by the prominent emotions athletes said they felt (confidence and fear), strategies that were tested in previous research (goal setting, imagery, relaxation, behaviour modelling), and what athletes said they needed (support to: set goals, manage pain and injury risk, understand their injury and what was required for physical and mental recovery, receive feedback on their recovery progress).

Internet-delivered psychological support works best when users feel engaged in their mental health support in real time, when the intervention employs a user-friendly interface that prioritises multimedia, and when the intervention structure encourages users to engage in self-monitoring.[23] We embraced the recommendations for designing mental eHealth interventions and established 5 key design principles (Highlight box 5).

### Highlight box 5

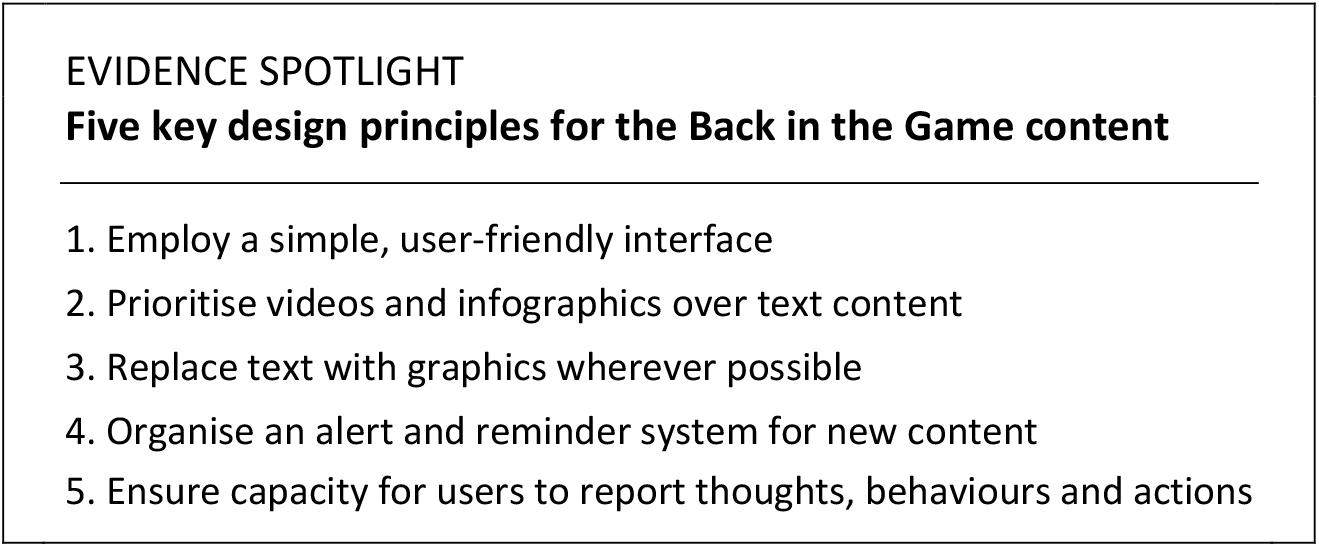

### Building block 8: Consider real-world issues

Back in the Game is self-directed psychological support intervention designed to be available on-demand and delivered via the Internet (smartphone app or website). An intervention that complements physical rehabilitation might be an effective way to overcome some of the geography, stigma and cost barriers to delivering effective psychological support to athletes during and after rehabilitation. eHealth technology facilitates low-cost, on-demand delivery of psychological support to injured athletes. Smartphones are an accessible platform from which to deliver evidence-based strategies for improving confidence to return to sport that athletes can access anywhere, and at any time.

For at least two decades, clinicians and health researchers have developed and delivered psychological treatments via the Internet.[4] Digital interventions can be as effective as visiting a psychologist in-person, deliver sustained benefits, and are probably cost effective.[3] Using digital interventions to complement face-to-face sports injury rehabilitation is an area of growing research interest.[26, 45, 63]

### Refining and validating Back in the Game content and design

After designing the first iteration of the intervention, we completed a two-phase feasibility testing project (the project was approved by the Swedish Ethical Review Authority (2018/41-31)). Because Back in the Game is a new intervention, we wanted to ensure it was appropriate and acceptable for athletes after ACL reconstruction. Detailed methods and results for the feasibility and usability testing are in Appendix 3.

In phase 1 of feasibility testing, we focused on addressing practical issues. We sought feedback regarding technical problems with the platform used to deliver and display content, not specifically regarding the intervention content. In phase 2, we invited feedback via multiple rounds of interviews on the intervention content, look and feel of the user interface, flow and acceptability of content, frequency of content delivery and value of the intervention.

We worked with the content delivery platform vendor (Briteback AB, Norrköping, Sweden) to complete additional engineering and implement bug fixes in response to feedback we received in phase 1 (Table A3a (Appendix 3) details the specific bugs and bug fixes). In phase 2, we identified aspects of the intervention that were satisfactory and did not require changes to the intervention (Table A3b), and aspects that required specific changes/actions (Table A3c).

### Summary of user feedback on Back in the Game

Users (i) thought the intervention would add value to their physical rehabilitation, (ii) found appealing content with appropriate flow, (iii) appreciated receiving notifications and reminders to engage with the intervention, and (iv) said the goal-setting module was helpful. Users wanted (i) more support to get started with the intervention, (ii) to better understand what they had to do, when and why, and (iii) more feedback on their progress through the intervention. In response to the feasibility testing results, we added additional video and infographic content, and strengthened the system of providing tailored progress reports and feedback.

## Perspectives

In outlining the steps we took to develop the Back in the Game intervention, we hope to provide a starting point for future intervention developers who are refining existing interventions and developing new ones. We describe a multi-faceted approach to development, which considered the athlete’s lived experience and context, previous work in the field, a theoretical rationale for the intervention, and input from end users to ensure an appropriate and acceptable final product.

Four characteristics define Back in the Game as a complex intervention[27]:

i. Different components that interact
ii. Requires different actions from those who receive the intervention
iii. Variability in outcomes
iv. Flexible and tailored to the user

We described our blended approach to developing the Back in the Game intervention, guided by O’Cathain’s et al. taxonomy and synthesis.[53] Our work extends beyond developing the intervention—we are currently conducting a multi-centre pragmatic randomised controlled trial[9] to evaluate the efficacy of the intervention, and will report the trial results separately.

### Lived experience, needs and perspectives of injured athletes

Athletes lack formal, systematic psychological support during rehabilitation and return to sport. Psychological responses are prominent, and athletes want support to help manage their psychological response to injury and return to sport. Yet there are few psychological support interventions for injured athletes in the published literature.[25] Rehabilitation clinicians recognise athletes’ need for psychological support, but are often unsure how to provide sufficient psychological support for return to sport.

### Scope of the problem

As many as 1 in every 2 athletes do not return to their preinjury sport after ACL reconstruction. For those who do return, the transition through rehabilitation and resuming sport can be difficult as athletes experience concerns about performing as before injury, and anxiety about sustaining a new injury. Fear of reinjury is the most common reason for not returning to sport.[10, 13]

### Context of the target population

Athletes who wish to return to high knee demand activities often require ACL reconstruction surgery. Post-operative rehabilitation takes at least 9 months before it is safe to return to unrestricted sport participation.

### Effect of psychological support interventions for injured athletes

Psychological skills training that targets coping strategies, relaxation and goal setting might improve physical impairments and psychological outcomes after ACL reconstruction. Previous psychological support interventions for athletes have focused on early rehabilitation, and have not had an explicit focus on improving return to sport outcomes—a defining feature of Back in the Game.

### Aim of Back in the Game

The intervention aims to deliver on-demand psychological support after ACL reconstruction. The goal is to improve athletes’ confidence to return to sport and the number of athletes who safely return to their preinjury sport after ACL reconstruction.

### Ways to foster return to sport behaviour change

Combining cognitive theory and self-determination theory, we propose that when an athlete thinks positively about returning to sport, their motivation to return to sport is enhanced. Back in the Game employs cognitive behavioural therapy and motivational interviewing principles to change negative thinking about returning to sport to positive thinking, and ultimately boost the athlete’s motivation to return to sport.

### Framework for Back in the Game

The intervention complements usual rehabilitation, and harnesses psychological skills, psychoeducation and motivational interviewing principles over a 24-week self-directed programme, delivered via Internet (smartphone app or web platform).

### Real world issues

Feedback we received from end users suggested that the Back in the Game intervention met our objective to develop a psychological support intervention that was easy-to-access, focused on return to sport, and complemented usual post-operative rehabilitation (i.e. the intervention was appropriate and acceptable for the target population). End users had a generally positive attitude to the intervention.

## Conclusion

The Back in the Game intervention is a 24-week Internet-delivered programme covering psychological skills, psychoeducation and principles of motivational interviewing. The self-guided intervention complements usual rehabilitation, changes focus as rehabilitation progresses, is easy to access and use, and includes different psychological support strategies.

## Data Availability

All data are included in the manuscript

## Acknowledgements

Thank you to Professor Nicholas Taylor who contributed to project planning, including designing feasibility studies and planning the efficacy trial, and to Professor Julian Feller who helped secure funding to support content design and production. Thank you to David Brohede, Linnéa Helfrich, Anna Wretman and Dr. Björn Paxling for help during content design and production; and to David Brohede and Anna Wretman for conducting the feasibility study interviews. Dr. Johan Åberg and the Briteback team provided technical support for the content delivery platform.

