## Appendix 1 for "Developing a psychological support intervention to help injured athletes get Back in the Game"

### APPENDIX 1. Understanding injured athletes' lived experience, perspectives and needs

In this section, we describe the methods and results addressing the question: *From the perspective of an active person with ACL reconstruction, in the setting of completing or having completed rehabilitation, how does the phenomenon of biopsychosocial factors during recovery impact on a person's experiences and perceptions related to recovery and return to sport?*

#### Search approach

We searched the MEDLINE, EMBASE AND SPORTDiscus electronic databases from inception (updated on 22 April 2020) using the key words "anterior cruciate ligament" AND ["interview" OR "qualitative"]. We deliberately used broad search terms because searching for qualitative syntheses is notoriously challenging.[53, 63] We supplemented the search with forward citation tracking (using Google Scholar), related paper searching, and hand searching the reference lists of included records.

#### Study selection

We selected studies that examined the opinions of people with ACL reconstruction about at least 1 issue related to returning to sport after ACL reconstruction, and that were reported in English language. We excluded studies that used mixed-methods, semi-quantitative or quantitative methods.

One researcher conducted the database search; a second researcher selected the articles. Records were exported to EndNote X9, and duplicates were removed before screening. Article selection occurred in two steps. First, the researcher screened the titles and abstracts of all records. Where it was unclear from the title and abstract whether an article should be included, the article was obtained in full text and screened. Once the final list of included articles was established, a second researcher checked the list and agreed on the final list.

#### Data extraction

One researcher extracted study and participant characteristics (sample size, sex, age, country, and population), and key results statements from each included study; a second reviewer cross-checked the accuracy of the data extraction. The key results statements were organised in a table ready for coding to commence.

#### Data synthesis

We used a thematic synthesis approach[58, 65] to gather, code and summarise information, and identify key themes and a novel interpretation of the body of qualitative literature[58, 65]. Coding was an iterative process through which we established and refined a thematic framework for synthesising information. The data extraction, coding and synthesis approach occurred in parallel, concurrently refining and developing the synthesis framework as themes emerged. Quotes from study participants and the descriptions of key study results informed new themes from the available data.

There were 4 steps in the synthesis process:

1. A researcher read each article and completed free line-by-line coding of the results, which was cross-checked by a second researcher.
2. Two researchers collaborated to establish new descriptive themes from the line-by-line coding.
3. One researcher wrote a summary of the synthesised results, organised by the 16 descriptive themes; a second researcher cross-checked the summary.
4. Two researchers distilled the descriptive themes to new analytic themes that addressed the aim of reviewing the literature.

We consistently reviewed and cross-checked the descriptive theme labels so that earlier coding could guide later coding, and we could update earlier coding as new information emerged. Finally, we reviewed all descriptive themes and wrote descriptive summaries based on the line-by-line coding. Two reviewers worked on the coding the synthesis: line-by-line coding and descriptive themes were identified by each reviewer independently; one reviewer wrote the descriptive summaries, and the second reviewer cross-checked the descriptive summaries for accuracy. We present the final results in summary tables.

#### Results

We identified 240 records. After deleting duplicate records and screening, there were 16 qualitative studies included (Figure A1) that reported the experiences and opinions of more than 164 participants (1 study did not report how many participants with ACL reconstruction were interviewed). The participants were mainly professional or non-professional athletes, predominately in their teenage or young adult years. There were 77 female participants and 85 male participants (2 studies did not report sex). Studies were conducted in high income countries in Australasia (*n* = 3), Europe (*n* = 8) and North America (*n* = 5) (Table A1a).

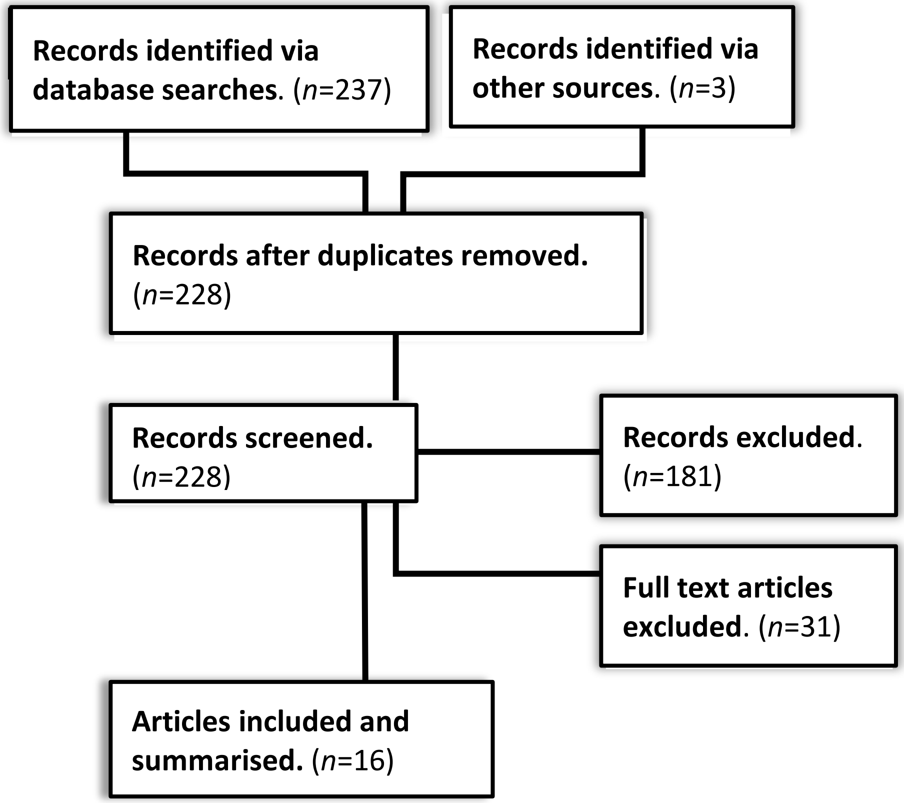

**Figure A1.** Identifying and selecting qualitative studies exploring the perceptions and experience of people with ACL reconstruction during rehabilitation and return to sport.

**Table A1a.** Summary of qualitative study characteristics

| Study identifier | Country | Population | n | Sex (F:M) | Age |
|---|---|---|---|---|---|
| Thing 2006[64] | Denmark | Non-professional handball players completing rehabilitation after ACL reconstruction | 17 | 17:0 | 19-33 years |
| Heijne 2008[40] | Sweden | Non-professional pivoting sport athletes, 12-21 months after ACL reconstruction | 10 | 1:9 | 23-41 years |
| Carson 2012[21] | England | Professional rugby union players during rehabilitation and transition back to sport | 5 | 0:5 | 18-27 years |
| Tjong 2013[67] | Canada | Recreational (*n* = 20), high school (*n* = 4), university (*n* = 6) and professional (*n* = 1) athletes at least 2 years after ACL reconstruction | 31 | 9:22 | 18-40 years |
| Nordahl 2014[50] | Sweden | Elite alpine skiers from a selective sports high school, 5 years after ACL reconstruction | 5 | 2:3 | 16-19 years |
| Gill 2016[35] | England | University students | NR | NR | NR |
| Johnson 2016[41] | Sweden | Elite football players, 4-9 months after a first-time ACL reconstruction | 8 | 13:0 | 25-35 years |
| Carson 2017[22] | England | Professional rugby union players during rehabilitation and transition back to sport | 5 | 0:5 | 18-27 years |
| Burland 2018[20] | United States | Athletes who were active in competitive sport before ACL injury, at least 1 year after ACL reconstruction | 12 | 6:6 | 16-44 years |
| Scott 2018[59] | New Zealand | Recreational pivoting sport athletes, 8-36 months after ACL reconstruction | 9 | 4:5 | 21-37 years |
| DiSanti 2018[29] | United States | High school pivoting sport athletes with ACL reconstruction who had not returned to sport | 10 | 7:3 | 15-18 years |
| Conti 2019[24] | Italy | Professional basketball players who had returned to basketball after ACL reconstruction | 10 | 0:10 | 22-36 years |
| Kunnen 2019[42] | Australia | Amateur (*n* = 17), national (*n* = 2) and international (*n* = 2) football players who had returned to sport | 21 | 9:12 | 19-51 years |
| Paterno 2019[54] | United States | Youth pivoting sport athletes who had and had not returned to sport | 10 | 4:6 | 11-19 years |
| Sole 2019[61] | New Zealand | Competitive pivoting sport athletes up to 5 years after ACL reconstruction | 9 | 5:4 | 18-26 years |
| Truong 2020[68] | Canada | Youth athletes up to 2 years after ACL injury, who had returned to sport | 7 | NR | 15-19 years |

*Note*. NR, not reported; [21] and [22] reported on the same sample.

#### Meta-synthesis

Three analytic themes emerged from the studies of athletes’ experiences, perceptions and needs, based on 16 descriptive themes (Table A1b):

1. Barriers and facilitators for *psychological readiness to return to sport*
2. Barriers and facilitators for *physical readiness to return to sport*
3. *Tools or strategies* to support rehabilitation progress

A psychological support intervention to help athletes return to sport after injury should include content targeting barriers and boosting facilitators for physical and mental readiness to return to sport, and aim to provide practical tools or strategies that athletes could use in parallel with their physical rehabilitation programme.

**Table A1b.** Athletes' perceptions and experiences related to recovery and return to sport after ACL reconstruction

| Descriptive theme (review finding) | Studies contributing to the review finding | Analytic theme |
|---|---|---|
| **Education** | | |
| Early in the rehabilitation period, athletes wanted to learn about what was required for successful outcomes after injury. They wanted to learn coping strategies that could help their recovery, including how to cope with pain and how to manage risk. Athletes saw the clinician as a guide to what they (the athlete) needed to do to reach their return to sport goals. | [22, 24, 35, 40, 54, 64, 68] | Tools or strategies |
| **Communication** | | |
| Constructive communication with others, including coaches and clinicians, helped athletes stay motivated and confident about their recovery and return to sport. Feeling uncertain hindered athletes' progress, and clinicians could help by clearly articulating the pathway and timeline for recovery. | [21, 29, 41, 59] | Tools or strategies |
| **Information** | | |
| Athletes wanted clear information about how long it would take to recover and return to sport. They also wanted information to help understand ACL injury and what rehabilitation would entail. When athletes understood their situation and what was required to recover, they felt more confident about the prognosis for recovery and return to sport. Having access to appropriate resources at the appropriate time during rehabilitation (e.g. gymnasium, clinicians, exercise programmes) was important, and athletes knew they needed to take proactive steps to ensure they stayed healthy after returning to sport. | [24, 29, 41, 54, 59, 68] | Tools or strategies |
| **Support from others/social support** | | |
| Athletes drew on social support from different places, including family, friends, rehabilitation clinicians, coaches, and teammates/peers. Athletes viewed social support as a critical part of their rehabilitation and vital for maintaining motivation and confidence to return to sport. Athletes drew support, feedback, encouragement and reassurance from people they trusted. | [20-22, 24, 29, 35, 41, 50, 54, 59, 61, 64, 68] | Tools or strategies |
| **Feedback** | | |
| Regularly measuring function and seeing it improve helped athletes stay motivated during rehabilitation. Athletes trusted rehabilitation clinicians to provide appropriate and timely feedback on rehabilitation progress. Athletes described receiving feedback as a key pillar of quality rehabilitation after ACL injury. | [22, 24, 40, 42, 54] | Tools or strategies |

| Descriptive theme (review finding) | Studies contributing to the review finding | Analytic theme |
|---|---|---|
| **Setting and achieving goals** | | |
| Setting goals helped athletes stay motivated and adequately prepare physically and mentally to return to sport. Some athletes had good support from clinicians to set rehabilitation and return to sport goals that mattered to them, while others received insufficient support. Athletes wanted support from clinicians to set goals, and saw goal setting and a structured approach to charting progress as a hallmark of quality rehabilitation. | [21, 22, 24, 29, 35, 40-42, 54] | Tools or strategies |
| **Coping strategies** | | |
| Coping with injury was facilitated by positive attitudes and personality traits (e.g. optimism, patience, determination) and cognitive processes like problem-solving, goal setting and rational thinking. Coping with injury was central to effectively completing rehabilitation. | [24, 50, 61] | Tools or strategies |
| **Staying injury free** | | |
| Fear about sustaining another ACL injury was one of the most prominent emotions for athletes during rehabilitation and when returning to sport. Accepting that there was risk inherent in playing sport, and learning how to manage that risk, including learning strategies to help prevent new ACL injuries, were important to athletes. Athletes recognised that they were responsible for continuing strategies like injury prevention exercises after returning to sport. Athletes said that good rehabilitation programmes included strategies to help build their physical and mental capacity to participate safely in sport. | [29, 35, 42, 59, 61, 64, 67, 68] | Barriers and facilitators for physical readiness to return to sport |
| **Capacity of one's body to perform in sport** | | |
| Athletes were nervous about whether they could perform well in their sport after returning from injury, and felt personal responsibility for their progress towards full recovery. Athletes felt confident about their rehabilitation preparing them for the demands of performing their sport, but they were concerned about sustaining another injury while playing sport. Performing sport-specific movements and tasks during rehabilitation, and using visualisation/mental imagery, helped athletes prepare psychologically to perform well when they returned to their sport. | [20, 21, 24, 29, 40, 42, 50, 67] | Barriers and facilitators for physical and psychological readiness to return to sport |
| **Emotions** | | |

| Descriptive theme (review finding) | Studies contributing to the review finding | Analytic theme |
|---|---|---|
| Athletes felt scared, uncertain, frustrated, and hopeless at different times during recovery. Sometimes they avoided activities because of fear or lack of confidence. Athletes' fear and anxiety was typically about getting the injury again, having to go through rehabilitation again, the long-term consequences of injury and whether they could perform well again. Athletes felt happy and free when playing their sport. | [20, 21, 29, 35, 40-42, 50, 54, 59, 64, 67, 68] | Barriers and facilitators for psychological readiness to return to sport |
| **Expectations** | | |
| Athletes expected to learn realistic timelines for recovery and return to sport from the clinician. Feeling uncertain about their progress hindered mental preparation for returning to sport, but when athletes understood the process, they felt confident and prepared. Athletes' expectations affected how ready they felt to contribute to decisions during rehabilitation and return to sport. | [20, 22, 24, 29, 35, 40, 54, 59] | Barriers and facilitators for psychological readiness to return to sport |
| **Motivation** | | |
| Receiving feedback, clear communication with and support from trusted people, setting and achieving goals, self-belief and a love of sport/playing sport boosted athletes' motivation. Athletes were typically motivated to return to sport, especially early in the recovery process. Although, losing patience during the long and monotonous rehabilitation depleted their motivation. | [20, 24, 29, 40-42, 50, 54, 59, 61, 64, 67, 68] | Barriers and facilitators for psychological readiness to return to sport |
| **Confidence** | | |
| Athletes lacked confidence at different stages during recovery from injury, and lacking confidence was a key barrier to returning to sport. When they lacked confidence, athletes responded by self-limiting participation in sport. Regaining confidence was the most important focus of recovery and returning to sport. Having strong support from their social network (including from rehabilitation clinicians) and seeing progress towards their goals during rehabilitation helped athletes build their confidence to return to sport. | [20, 21, 24, 42, 50, 54, 59] | Barriers and facilitators for psychological readiness to return to sport |
| **Self-efficacy for recovery from injury** | | |
| During recovery from injury and rehabilitation many athletes lacked self-efficacy and self-esteem. If athletes had previous experience of injury or were an experienced and established member of their team, they were able to draw on these experiences to build resilience during recovery. Previous experience helped athletes know what they needed to do to recover from injury. | [40, 41, 50] | Barriers and facilitators for psychological |

| Descriptive theme (review finding) | Studies contributing to the review finding | Analytic theme |
|---|---|---|
| | | readiness to return to sport |
| **Identity as an athlete and as a member of society** | | |
| Athletes saw themselves as athletes when they were injured, not as patients. Athletes wanted to play sport and felt that playing sport was a central part of who they were as a person. Many athletes had strong self-belief for returning to sport, but were also nervous about their capacity to perform well after returning. For some athletes, their experience of injury had irrevocably changed how they thought of themselves and their place in and contribution to society. Identifying as an athlete was a strong motivating factor for return to sport. | [20-22, 41, 42, 50, 59, 64, 67] | Barriers and facilitators for psychological readiness to return to sport |
| **Priorities for playing sport** | | |
| After experiencing a knee injury, athletes spent time thinking about their preferences and priorities about participating in sport. Athletes recognised that being regularly physically active was beneficial for health. Athletes were aware that there were risks associated with returning to sport after ACL injury. Some athletes decided that the risk for sustaining a new knee injury was not worth the benefit of returning to play their previous sport; other athletes accepted the risk and the need to take active steps to manage risk after returning to their sport. | [20, 59, 61, 67, 68] | Barriers and facilitators for psychological readiness to return to sport |
