## Appendix 2 for "Developing a psychological support intervention to help injured athletes get Back in the Game"

### APPENDIX 2. The state of play: credible causal explanation for a psychological support intervention after ACL reconstruction

Previous reviews[25, 34] did not include grey literature, and included study designs other than randomised controlled trials (RCTs). Given the growing interest in the field, and because we knew there were few published interventions focused on improving return to sport outcomes, it was important to include grey literature. Given we aimed to build a credible causal case for a psychological support intervention, it was important to limit the literature search to designs that could answer a causal question (i.e. RCTs).

#### Search approach

We searched the electronic databases PsycINFO, MEDLINE, SPORTDiscus, CINAHL, EMBASE, PubMed and PsycARTICLES from database inception (updated on 21 May 2020; see Summary Box 1 for search strategy as applied to PubMed). We hand-searched the reference lists of included articles and previous reviews ([25, 34]) to identify articles not listed in the electronic database search, and searched clinical trial registries (ClinicalTrials.gov, EU Clinical Trials Register, ISRCTN Registry, and Australian New Zealand Clinical Trials Registry) on 22 May 2020, to identify any relevant unpublished trials.

***Summary Box 1.*** *Search strategy as applied to PubMed for literature search to address the question "What is the efficacy of psychological interventions for improving ACL injury rehabilitation outcomes in athletes?"*

((((((Psychological Intervention OR ((Cognitive-behavioral therapy) OR (Cognitive) OR (Relaxation) OR (Imagery) OR (Guided Imagery) OR (Acceptance and Commitment Therapy)) OR Psychological Therapy OR (Behavioral Intervention AND (Physical Therapy)) OR (Therapeutics AND ((Mind-Body Therapies [MeSH]) OR (Acupuncture) OR (Message) OR (Goal setting) OR (Psychological Skills) OR (Meditation) OR (Videoinsight) OR (Psychotherapeutic)))))) AND ((rehabilitation outcome[MeSH] OR patient relevant outcomes OR (functional improvement) OR (strength) OR (anxiety) OR (return to sport) OR (functional outcome) OR clinical effectiveness OR Knee Outcomes))) AND (Anterior Cruciate Ligament Reconstruction[MeSH] OR ACL Reconstruction OR ACL OR ACL Surgery OR Anterior Cruciate Ligament Surgery)))

#### Trial selection

We selected randomised controlled trials of psychological interventions administered before or after ACL reconstruction. To be included, trials had to fulfil the following criteria:

**Population.** At least 50% of participants who completed final follow-up were skeletally mature athletes (any level) with ACL reconstruction

**Intervention.** Any treatment aimed at addressing a psychological response/factor (e.g. cognition or emotion)

**Comparison.** Any reasonable comparison (e.g. usual rehabilitation care)

**Outcome.** Measured at least 1 rehabilitation-related outcome (e.g. self-reported knee function, muscle strength, return to sport rate, psychological response during treatment)

One reviewer conducted the database search and selected the articles for inclusion. First, the reviewer screened the titles and abstracts of all records. Where it was unclear from the title and abstract whether an article should be included, we obtained and screened the article full text. We contacted authors to clarify information related to eligibility screening, as required.

#### Data extraction

One reviewer extracted the following data elements from each included article: population (including number of participants), details of the psychological intervention, comparison, primary outcome(s) and primary trial end point. A second reviewer checked the data extraction for accuracy.

For an estimate of the effect of each intervention, one reviewer extracted the effect estimate, or sufficient data (e.g. mean, standard deviation, sample size) to calculate an effect estimate for the primary outcome(s) at the primary time point in each trial. Where multiple follow-ups were reported, we prioritised the longest follow-up from baseline. Where multiple primary outcomes were reported at the same time point, we extracted data for each outcome.

For an overview of trial quality, we extracted the Physiotherapy Evidence Database (PEDro) score, where available, from the PEDro website (www.pedro.org.au). If the trial was not indexed in the PEDro database, one reviewer assigned a PEDro score.[49]

#### Data summary approach

We identified 529 records. After deleting duplicate records, and screening, there were 7 trials to summarise (Figure A2). We summarised the trial data elements, PEDro scores and effect estimates (Table A2), then produced descriptive summaries of the treatment approaches that had been studied. We used RevMan Version 5.3 (Copenhagen: The Nordic Cochrane Centre, The Cochrane Collaboration, 2014) to calculate mean difference and 95% confidence intervals for primary outcomes. A second reviewer checked the effect estimate calculations.

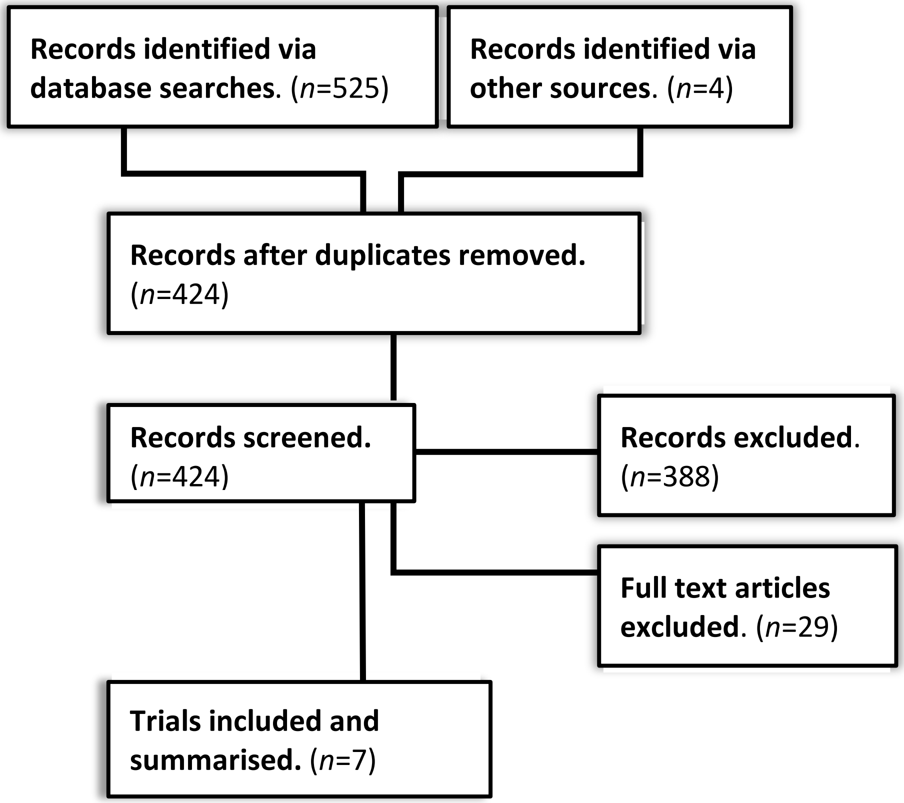


**Figure A2.** Identifying and selecting psychological interventions in ACL rehabilitation

#### Descriptive summary of results from 7 randomised controlled trials

The effect of sports psychology interventions on improving rehabilitation outcomes after ACL reconstruction have been studied for at least two decades. The most extensively studied psychological support interventions in sports rehabilitation were imagery, relaxation and goal setting.

##### Imagery or visualisation

Athletes completed up to 10 sessions where they mentally rehearsed the specific skills or movements they were executing in physiotherapy rehabilitation, and imagined their body moving through stages of physiological healing.[28, 48] There was conflicting evidence regarding the effects of imagery training on quadriceps strength, reinjury anxiety and self-efficacy.

##### Relaxation

Combining breath-assisted relaxation with guided imagery was effective for reducing pain and reinjury anxiety, and improving quadriceps strength in athletes at 6 months after ACL reconstruction compared to placebo or control.[28] Because it was a combined intervention, it was unclear whether relaxation alone might have yielded similar results.

##### Goal-setting

In a five-week goal setting intervention conducted by a sport psychologist in parallel with physiotherapy rehabilitation, athletes spent 60 to 105 minutes per week setting rehabilitation goals.[32] Athletes also completed a daily diary to document their progress and emotions during recovery. Goal setting might improve rehabilitation adherence and self-efficacy compared to social support or control.[32]

##### Coping modelling

Participants watched a video of people with ACL injury demonstrating (modelling) coping strategies including rehabilitation exercises and sharing their thoughts, feelings and rehabilitation progress. Participants who watched the videos had superior self-reported knee function and early rehabilitation self-efficacy compared to control.[47]

#### Therapeutic insights

After watching a contemporary art video for 2 months in combination with usual postoperative rehabilitation, participants who received the video designed to elicit positive therapeutic insights had superior self-reported knee function compared to participants who watched the video designed to elicit unfavourable insights.[73]

**Table A2.** Summary of psychological interventions for improving rehabilitation outcomes after ACL reconstruction

| Trial identifier | Population | Intervention details | Comparison | Primary outcome(s) | Primary endpoint(s) | Effect estimate | PEDro score |
|---|---|---|---|---|---|---|---|
| Cupal 2001[28] | 30 athletes (recreation or competitive level) with ACL reconstruction | 10 individual **guided imagery** and **relaxation** sessions completed 2 weeks apart<br>**Mental rehearsal** of rehabilitation goals<br>Suggestions to promote positive **emotional coping** responses | Placebo imagery: instructions to spend 10-15 minutes per day sitting quietly and visualising a peaceful scene | Knee strength (muscle groups tested were not defined)<br><br>Re-injury anxiety<br><br>Physical pain | 6 months post-operative | MD: 0.17 (0.06 to 0.28)<br><br>MD: 2.3 (1.4 to 3.2) VAS points<br><br>MD: 2.0 (1.1 to 2.9) VAS points | 5 |
| Evans & Hardy 2002[32] | 30 athletes (recreation or competitive level) with ACL or PCL surgery out of a total of 39 participants recruited from a sports injury clinic | **Goal-setting** for rehabilitation tasks and progress (5-week programme, 1 × 60-105 minute session each week)<br>Daily diary to document progress and emotions | Social support: emotional support from a sport psychologist (40-60 minute sessions every 7-10 days for 5 weeks), plus daily diary to document progress and emotions.<br><br>Control: Telephone call from a sport psychologist every 10 days to encourage | Rehabilitation adherence<br><br>Self-efficacy<br><br>Treatment efficacy<br><br>Dispirited psychological | 5-weeks after starting the intervention (patients were at different post-operative timepoints) | GS vs. C: MD 28.3 (13.9 to 42.6)<br><br>GS vs. C: 7.0 (4.5 to 9.6)<br><br>GS vs. C: 4.5 (2.0 to 6.9) | 4 |

| | | | | | | | |
|---|---|---|---|---|---|---|---|
| | | | adherence to the study. | response to injury<br><br>Reorganisation psychological response to injury | | GS vs. C: MD -1.7 (-3.8 to 0.4)<br><br>GS vs. C: MD 3.4 (1.6 to 5.2) | |
| Maddison 2006[47] | 58 patients with ACL reconstruction completed final follow-up | Two **coping modelling** videos. Video 1 was viewed pre-operatively and at discharge from hospital. Video 2 was viewed at 2- and 6-weeks post-operative. The videos comprised interviews with previously injured athletes, and still images of athletes completing rehabilitation tasks | Usual rehabilitation care | Self-reported knee function (IKDC subjective knee form)<br><br>Walking self-efficacy<br><br>Exercise self-efficacy | 6 weeks post-operative | MD: 4.2 (0.4 to 8.7) points<br><br>MD: 3.7 (1.4 to 8.8)<br><br>MD: 3.2 (3.1 to 9.5) | 4 |
| Maddison 2012[48] | 21 participants, aged at least 16 years, with primary ACL reconstruction | 9 individual sessions of **guided imagery** and **relaxation**.<br>**Mental rehearsal** of activities to achieve ACL rehabilitation goals | Usual rehabilitation care | Knee extension strength (60°/s) | 6 months post-operative | MD: 11.5 (-16.1 to 39.1) Nm | 7 |
| Zaffagnini 2013[73] | 101 of 106 participants, aged at least 16 years, with primary ACL reconstruction | Watching an art video with images designed to induce **positive and therapeutic insights** into one's psychological experience— | Watching an art video with images designed to induce psychological insights unfavourable | Self-reported knee function (IKDC subjective knee form) | 3 months post-operative | MD: 11.0 (4.4 to 17.7) points | 9 |

| | | | | | | | |
|---|---|---|---|---|---|---|---|
| | completed final follow-up | to promote learning, emotions, physical and mental recovery. Instructions to watch the video 3 times per week for the first 2 post-operative months | to psychological recovery. Instructions to watch the video 3 times per week for the first 2 post-operative months. | | | | |
| Archer 2020 (NCT 03243162) | 90 participants, aged 14 to 35 years, who were active in sport prior to ACL injury | **Cognitive-behavioural-based physical therapy** (7-session programme delivered via telephone over an 8-week period—1 pre-operative telephone session; 6 post-operative telephone sessions), addressing recovery expectations, fear of movement or reinjury, self-efficacy, and coping | Education programme focused on post-operative recovery (7-session programme delivered via telephone over an 8-week period—1 pre-operative telephone session; 6 post-operative telephone sessions) | KOOS sport & recreation subscale score<br><br>Marx Activity Scale score | 12 months post-operative | N/A | N/A |
| Ageberg 2020 (NCT 03473821) | 100 participants, aged 18 to 35 years, who were active in competitive or recreational sport prior to ACL injury | **Dynamic motor imagery** combined with neuromuscular training. In dynamic motor imagery, the person imagines performing a rehabilitation task | Usual rehabilitation care | Side-hop performance<br><br>ACL-Return to Sport after Injury scale score | 12-weeks post-operative<br>12 months post-operative | N/A | N/A |

*Note.* MD, mean difference; GS, goal-setting group; C, control group
