## Appendix 3 for "Developing a psychological support intervention to help injured athletes get Back in the Game"

### APPENDIX 3. Feasibility and useability of the Back in the Game intervention

Feasibility testing has different applications in the context of developing an intervention. The underlying motive is to establish whether it is appropriate to continue studying an intervention. Researchers considering feasibility studies are aiming to establish whether ongoing research will be worthwhile and sustainable. There are at least 5 scenarios within which researchers may consider conducting a feasibility study:[18]

(i) To establish, or deepen community partnerships
(ii) An intervention has not been tested before
(iii) It is unclear whether an existing intervention is appropriate for a specific population
(iv) An existing intervention needs tailoring to suit a specific population
(v) An existing intervention has not been effective for a specific population, but it is possible that an improved version of the intervention might have benefits

Back in the Game had not been tested before, and we wanted to ensure it was appropriate and acceptable for athletes with ACL reconstruction. Therefore, we evaluated the design and delivery of the intervention in two phases, conducted between July 2018 and May 2019. Phase 1 was a technical assessment; phase 2 was formal feasibility and useability testing.

#### Design

In phase 1 (technical assessment), we invited clinicians, researchers, and people with lived experience of sports injury to use the intervention and provide feedback about its basic functions. We focused on addressing practical issues related to the processes of creating an account and signing-on to access the intervention content, and how content was delivered and displayed. We asked for feedback regarding technical problems with the platform used to deliver and display content, not specifically regarding the intervention content.

In phase 2 (feasibility and useability testing), we invited patients who represented the intervention target population to participate in a field-based test of the intervention. The aim was to evaluate 4 aspects of feasibility[18]—*acceptability*, *demand*, *practicality* and *integration*; we simultaneously assessed *useability*. We invited feedback on the intervention content, look and feel of the user interface, flow and acceptability/appropriateness of content, frequency of content delivery, and value of the intervention. We did not evaluate efficacy (e.g. pilot case-control study to collect data regarding efficacy of the intervention) because we aimed to identify refinements and improvements to make to the intervention prior to completing a formal efficacy trial.[9]

The combined feasibility and field-based useability study was approved by the Swedish Ethical Review Authority (2018/45-31). Participants provided written, informed consent to participate, and the study was conducted according to the principles of the Declaration of Helsinki. We used a descriptive approach, conducting individual semi-structured interviews to evaluate *acceptability*, *demand*, *practicality*, and *useability* (Summary Box 2), and pre-post analysis of user statistics to evaluate *integration*.

#### Participants

For phase 1, we used strategic sampling to identify participants via clinical and professional networks. We recruited physiotherapists, orthopaedic surgeons, sports psychologists, researchers from the field of musculoskeletal rehabilitation, and people with lived experience of ACL injury.

For phase 2, we used strategic sampling to recruit patients with ACL reconstruction. Our sampling approach ensured we recruited participants who would provide constructive information that we could take action on, if required, and was appropriate to address the aims of the study. We recruited people who were aged between 15 and 30 years at the time of ACL injury and had sought treatment for ACL injury at one of two centres in south-eastern Sweden—a metropolitan private hospital, and a University hospital. Participants must have had primary ACL reconstruction no more than 8 weeks before starting the feasibility study, were fluent in written and spoken Swedish language, regularly participated in sport that involved pivoting and/or cutting (e.g. football, basketball, floorball) prior to ACL injury, and intended to return to their sport.[9] We aimed for the selection criteria to match the criteria we planned to use in our definitive efficacy trial as much as possible, while retaining a pragmatic approach and ensuring timely completion of the feasibility study.[9]

*Summary Box 2. Semi-structured interview guide*

What was it like to get started using the app?

What can we do to help you remember to answer questions/access content etc.?

How do you perceive the notifications?

What happens between you receiving a notification and doing something in the app?

What is your opinion of the flow of content in the app?

Were there parts that you found more difficult or easier to understand? Can you describe them?

Do you have suggestions for how the information/workflow in the app can be made clearer and simpler, or improved?

In what ways do you think you can be helped by an app like Back in the Game? (value)

Would you recommend a friend/teammate with an ACL injury to use this app? Why/why not?

What is most annoying about the app?

If the app cost you 100 SEK/month to use, what would make if feel worth the cost?

Rank the content in the order you think was most helpful (most helpful at top of list)
☐ Video
☐ Text
☐ Audio
☐ Questions about confidence

Intervention

Back in the Game is designed to be delivered via Internet over a 24-week period as an addition to usual care post-operative rehabilitation following ACL reconstruction.[9] For phase 2 (feasibility and useability study), we condensed the intervention to a 10-week programme (Figure A3a) as a pragmatic approach to completing the study and not unnecessarily delaying the start of the efficacy trial. The intervention component of the feasibility and useability study was delivered over a 4-month period.

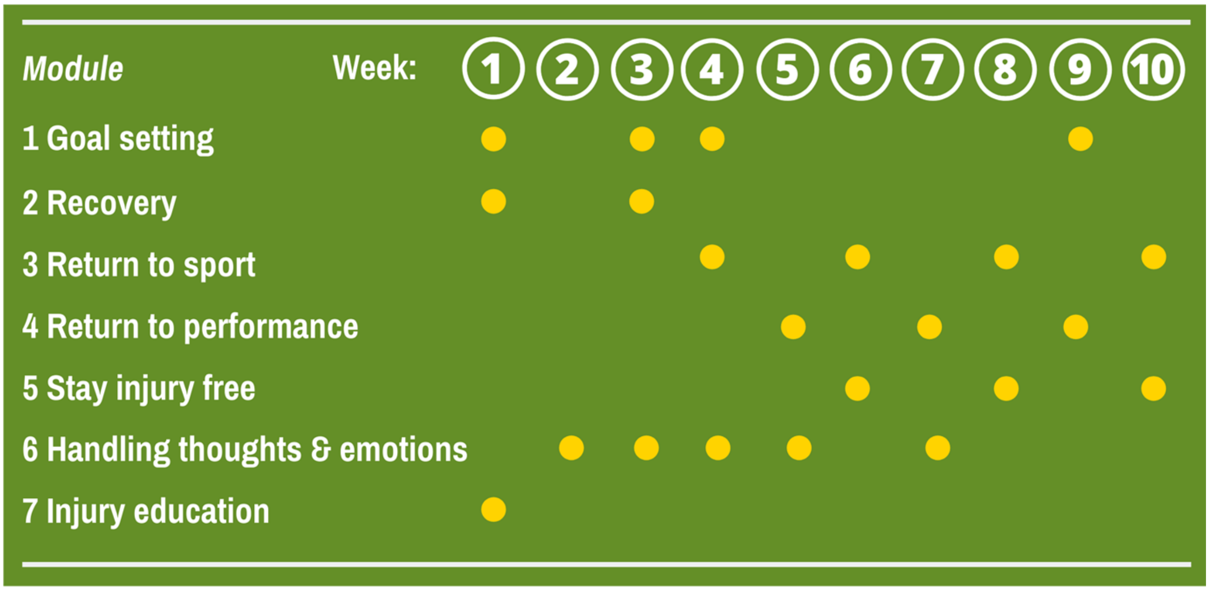


**Figure A3a.** Outline of content delivery over the 10-week Back in the Game programme. The programme comprises 7 self-directed content modules; timing of content delivery for each module is represented by a yellow dot.

#### Procedure

For phase 1 (technical assessment), the testers received an introduction email explaining the purpose, and with information about how to download the Briteback app (Briteback AB, Norrköping, Sweden; the platform we used to deliver the intervention content is available from the Apple App Store and Google Play Store), create a user account and get started. Briteback is a corporate communications software platform with an add-on content management feature for constructing and sending written, video and audio content, and administering questionnaires. The testers had access to all intervention content, and were encouraged to access the content as often as desired. We also asked the testers to ensure they accessed the content for each module at least once. Testers provided real-time feedback on the technical aspects of the intervention via telephone, email and direct chat within the Briteback platform.

For phase 2 (feasibility and useability study), participants received written information about the study, and were contacted by telephone approximately 1 week later, to determine eligibility. Participants who gave informed consent to participate received an email with information about how to download the Briteback app, create a user account and get started with the intervention. Participants completed a baseline questionnaire electronically, at least 1 day prior to receiving any intervention content. The questionnaire collected demographic characteristics, ratings of motivation to return to preinjury sport, self-efficacy (general self-efficacy scale[46]) and self-reported knee function (International Knee Documentation Committee subjective knee form[2]). Two licenced psychologists with experience developing and implementing Internet-delivered health interventions conducted one-on-one semi-structured interviews via telephone. Up to 4 interviews were conducted with each participant over the duration of the feasibility and useability testing period.

##### Evaluation approach/analysis

We used quasi-deductive content analysis to synthesise interview data. Each interview was transcribed and coded in an Excel spreadsheet. Coding and synthesis occurred in parallel. Two researchers collaborated to map and summarise participant feedback about intervention content, access and value. Statements from participants and the descriptions of key study results informed new themes from the available data. We summarise and describe the key feedback themes in summary tables.

We calculated descriptive statistics for the demographic characteristics and patient-reported outcomes (general self-efficacy, self-reported knee function) collected at baseline. We collated use statistics (total watch time for video, number of listens for audio) and completion rates for the intervention content. Participants also ranked the multimedia content types (video, text, audio, self-reflection) from most to least helpful (1 = most helpful; 4 = least helpful).

#### Results

Eleven people participated in phase 1 (technical assessment). Users identified 4 types of software errors (bugs) (Table A3a) that needed to be addressed prior to phase 2 testing. We worked with the software vendor (Briteback AB) to complete additional engineering to implement bug fixes and ensure the intervention (delivery platform and content) worked as intended prior to commencing phase 2. We also corrected typographical errors in the written content.

**Table A3a.** Summary of user feedback and actions from the technical assessment phase of Back in the Game intervention feasibility testing

| Software bug | Bug fix |
|---|---|
| Some user account names were visible to other users on the Briteback platform 'home screen' | Re-engineered the 'guest' user account type to ensure 'guest' accounts were masked to all other guest accounts |
| Informed consent page displayed twice to new users when they created an account | Re-engineered the consent process so new users only provided informed consent once upon joining the platform |
| Broken hyperlinks in audio and video content | Fixed all broken links and tested to ensure content displayed as intended |
| Some content that was set to automatically display after a set amount of time, either (i) did not display, or (ii) was displayed at an incorrect time | Re-engineered the automatic content display feature |

We invited 18 patients to participate in the feasibility and useability study. Five patients declined to participate; 6 did not reply. Seven participants with recent history of ACL reconstruction consented to participate in the combined feasibility and useability testing. Finally, 6 participants (3 women; 3 men) participated in the 10-week intervention and semi-structured interviews (Table A3b).

**Table A3b.** Demographic characteristics and motivation to return to sport, self-efficacy and self-reported knee function (data missing for one participant)

| Code | Work | Preinjury sport | Motivation (0-10) | | | GSES | IKDC |
|---|---|---|---|---|---|---|---|
| | | | Important[a] | Possible[b] | Invest[c] | (10-40) | (0-100) |
| O1 | Supervisor | Rugby | 9 | 10 | 10 | 30 | 68 |
| G2 | Doctor | Aerobics | 7 | 10 | 8 | 27 | 68 |
| P3 | Office worker | Football | 7 | 10 | 10 | 30 | 87 |
| M4 | Student | Handball | 9 | 10 | 9 | 39 | 72 |
| A5 | Project manager | Football | 9 | 8 | 8 | 31 | 66 |

[a] How important is it for you to return to your preinjury sport?

[b] Do you think it is possible for you to return to your preinjury sport?

[c] How much time and effort are you willing to invest to return to your preinjury sport?

GSES, general self-efficacy scale (higher score indicates greater self-efficacy); IKDC, International Knee Documentation Committee subjective knee form (higher score indicates superior self-reported knee function); motivation rated from 0 = not at all, to 10 = extremely.

Use statistics for the intervention were variable (Table A3c). Participants completed 14% to 86% of the intervention content during the feasibility study period. The content was available on demand, and we instructed and encouraged users to access the content they thought would help best when they felt they needed it. Given our instructions to users, and the framing of content as self-directed and available on demand, we did not expect participants would complete 100% of the intervention content.

Different types of multimedia content were helpful for different people (Table A3c). This supported our decision to employ a range of content types in the intervention, reflecting that different users have different needs, and that the intervention must allow the individual user to choose what to access and when.

**Table A3c.** Use statistics and participant ratings for multimedia content

| | **Video: total watch time (mins:sec)** | **Audio: listens (*n*)** | | | |
|---|---|---|---|---|---|
| **Month 1** | 42:30 | 6 | | | |
| **Month 2** | 24:15 | 4 | | | |
| **Month 3** | 48:00 | 5 | | | |
| **Month 4** | 9:00 | 3 | | | |
| **Content helpfulness rank** | **Video** | **Audio** | **Text** | **Self-reflection** | **Content completion** |
| **Participant O1** | 4 | 1 | 2 | 3 | 86% |
| **Participant G2*** | 1 | | | | 27% |
| **Participant P3** | 2 | 4 | 1 | 3 | 26% |
| **Participant M4** | 3 | 4 | 1 | 2 | 21% |
| **Participant A5^** | | | | | 14% |
| **Participant J7** | 1 | 3 | 4 | 2 | 19% |

*Note*. Helpfulness ranked from 1 = most helpful to 4 = least helpful; ^missing data; *participant G2 did not rate the helpfulness of the audio, text and self-reflection content.

Tables A3d and A3e outline synthesis of key feedback from participant interviews. Table A3d outlines participants' thoughts about the intervention and key components related to its delivery and content. Participants shared specific feedback about getting started, navigating intervention content and receiving real-time feedback on progress. The feedback guided additional content development and refined how existing intervention content was presented (Table A3e).

**Table A3d.** Feedback from participants that did not require action

| **Value of the intervention** |
|---|
| The app was helpful |
| The app improved motivation to participate in rehabilitation/physiotherapy |
| The app helped the user see that full recovery was achievable/to see a way ahead |
| Content helped the user to avoid being stuck worrying about the unknown |
| **Notifications** |
| A desired feature and a good reminder to prioritise working with the intervention content |
| **Content** |
| Getting started with the content was easy |
| The content flow was good |
| The content was appealing, especially the visualisation exercises, interviews with athletes and fear avoidance exercises |
| **Goal setting** |
| It was helpful to receive guidance for goal setting |
| Users enjoyed working on the goal-setting module |

**Table A3e.** Feedback that required action, and how we refined the intervention

| Feedback | Action |
|---|---|
| **Getting started—'onboarding'** | |
| Additional guidance on how to register a Briteback account and access Back in the Game would help participants know what to do. | Recorded an 'onboarding' video with step-by-step instructions of how to download and set up the app, and register with Briteback. Participants receive an 'onboarding' email with the link to the video plus additional written information about how to get set up with Briteback. |
| **Navigation and content flow** | |
| An overview of the full intervention would help users understand the time commitment for participating. | Recorded a content overview video (Figure A3b, Figure A3c) explaining the content, intervention structure and suggested time commitment, to complement the intervention introduction video. |
| It was difficult to understand the content arrangement. | 1. Produced an infographic with week-by-week breakdown of intervention content to complement the intervention introduction video.<br>2. Organised related content into separate content pages within the intervention home screen. |
| Some content seemed repetitious. | 1. Recorded a video explaining how the exercises are tailored to each stage of rehabilitation, and why some content repeats.<br>2. Made additional content available from the intervention home screen (i.e. easier to access on-demand). We organised the content into channels so users could easily find the desired content. |
| **User receiving feedback on progress** | |
| Users wanted more information about their progress through the intervention. | 1. Provided fortnightly progress reports outlining how many weeks of the intervention were completed and how many remain.<br>2. Feedback at the end of each module task outlining how many tasks were completed and how many tasks remain in the module.<br>3. |
| It was difficult to quickly find the previous goals set and to know whether the goals had been achieved (desire for more feedback about goals) | Personalised feedback screen for goals set. New goals are recorded on this screen; users must keep their own record of progress toward the goal(s) |

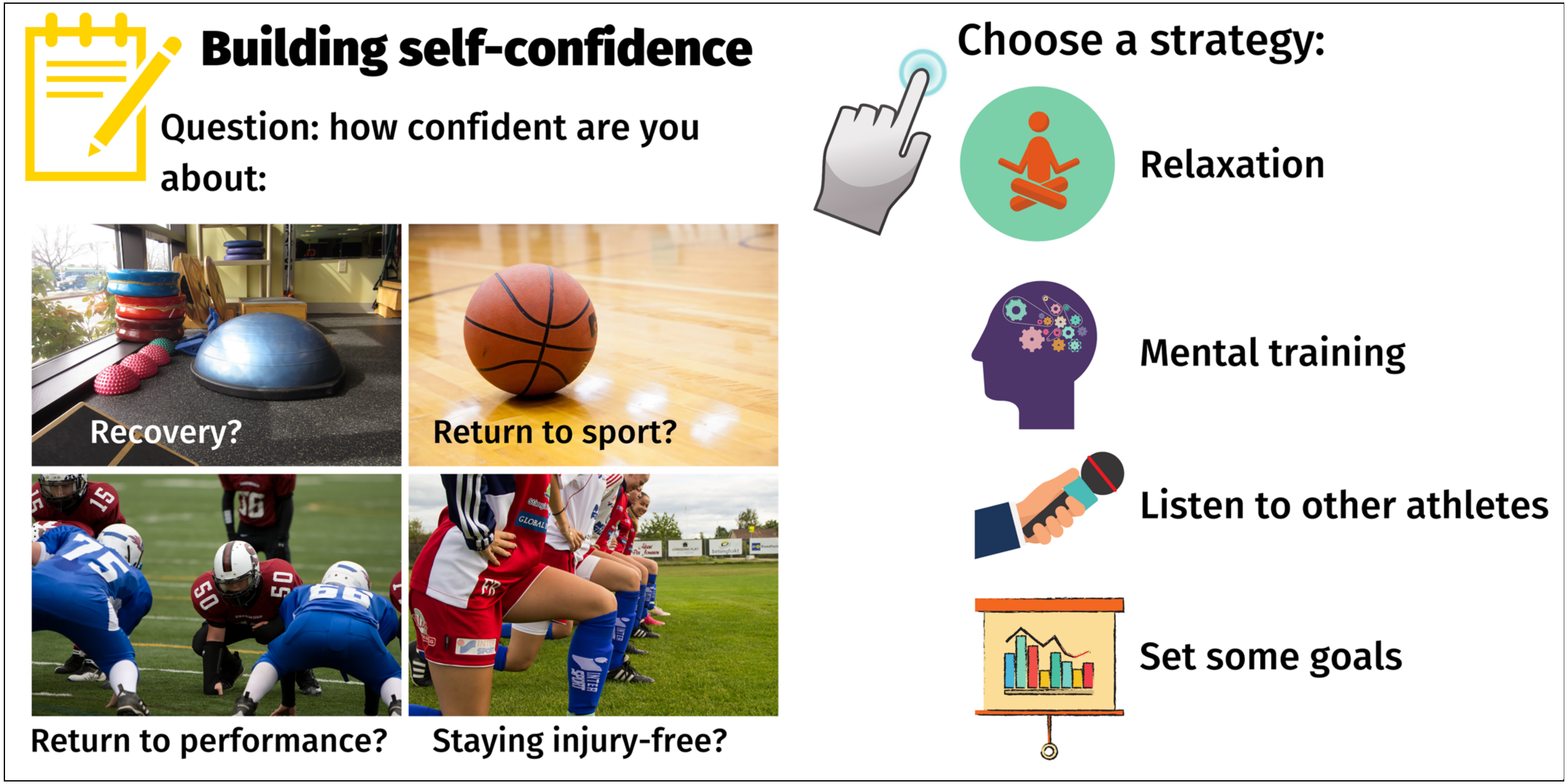


**Figure A3b.** Overview of the *recovery*, *return to sport*, *return to performance*, and *stay injury free* modules plus linked cognitive behavioural therapy tasks (strategies) [screenshot; as presented in the content overview video].

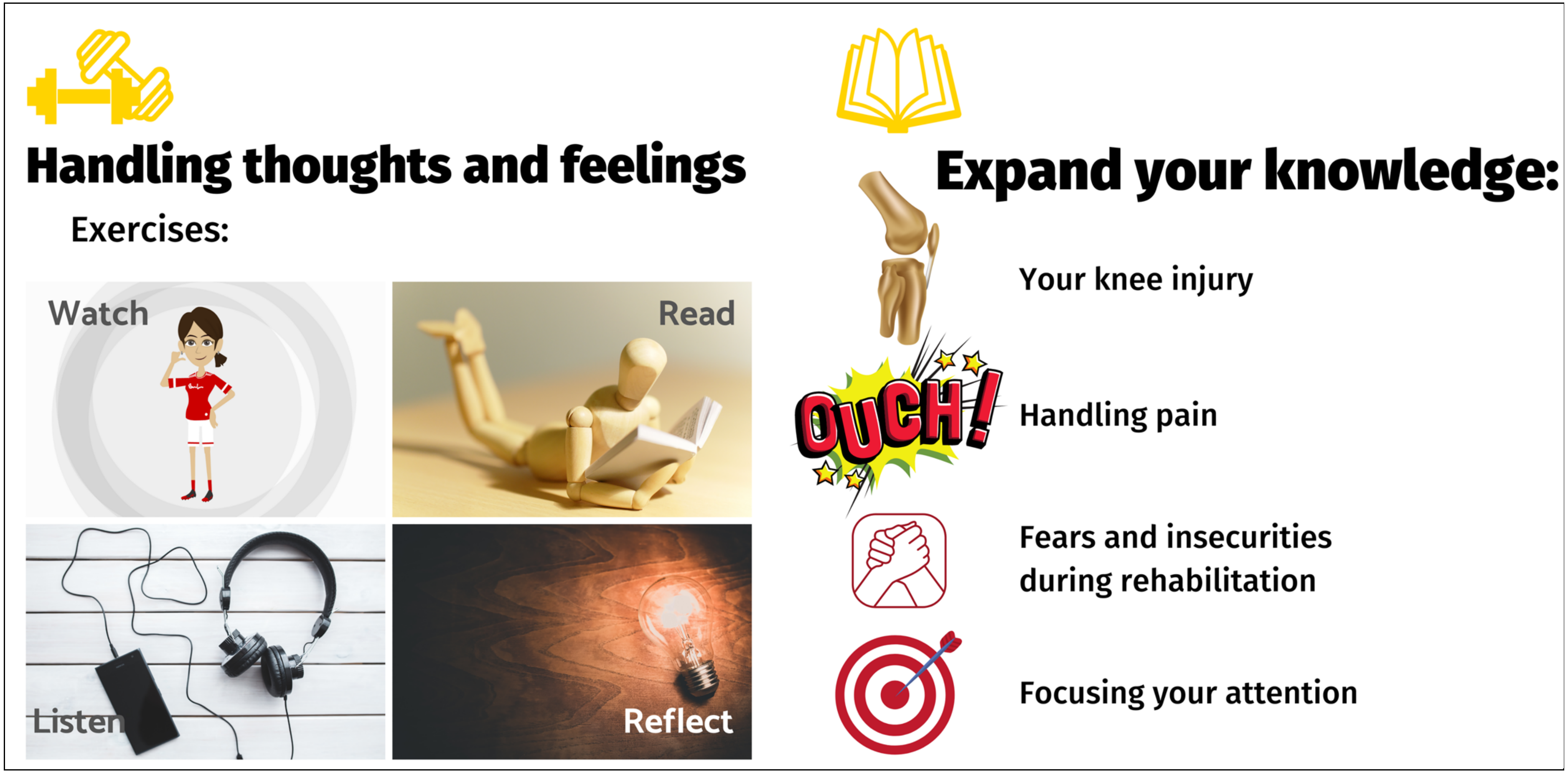


Figure A3c. Overview of the *handling thoughts & emotions*, and *injury education* modules [screenshot; as presented in the content overview video].
